# Trends in Mail-Order Prescription Use among U.S. Adults from 1996 to 2018: A Nationally Representative Repeated Cross-Sectional Study

**DOI:** 10.1101/2020.09.22.20199505

**Authors:** Duy Do, Pascal Geldsetzer

**Author notes:** (corresponding author) **Duy Do, PhD**.

## Abstract

**Background:** Mail-order prescriptions are popular in the U.S., but the recent mail delays due to operational changes at the United States Postal Services (USPS) may postpone the delivery of vital medications. Despite growing recognition of the health and economic effects of a postal crisis on mail-order pharmacy consumers, little is known about the extent of mail-order prescription use, and – most importantly – the population groups and types of medications that will likely be most affected by these postal delays.

**Methods:** The prevalence of mail-order prescription use was assessed using a nationally representative repeated cross-sectional survey (the Medical Expenditure Panel Survey) carried out among adults aged 18 and older in each year from 1996 to 2018. We stratified use of mail-order prescription by socio-demographic and health characteristics. Additionally, we calculated which prescription medications were most prevalent among all mailed medications, and for which medications users were most likely to opt for mail-order prescription.

**Findings:** 500,217 adults participated in the survey. Between 1996 and 2018, the prevalence of using at least one mail-order prescription in a year among U.S. adults was 9·8% (95% CI, 9·5%-10·0%). Each user purchased a mean of 19.4 (95% CI, 19·0-19·8) mail-order prescriptions annually. The prevalence of use increased from 6·9% (95% CI, 6·4%-7·5%) in 1996 to 10·3% (95% CI, 9·7%-10·9%) in 2018, and the mean annual number of mail-order prescriptions per user increased from 10·7 (95% CI, 9·8-11·7) to 20·5 (95% CI, 19·3-21·7) over the same period. Use of mail-order prescription in 2018 was common among adults aged 65 and older (23·9% [95% CI, 22·3%-25·4%]), non-Hispanic whites (13·6% [95% CI, 12·8%-14·5%]), married adults (12·7% [95% CI, 11·8%-13·6%]), college graduates (12·2% [95% CI, 11·3%-13·1%]), high-income adults (12·6%, [95% CI, 11·6%-13·6%]), disabled adults (19·3% [95% CI, 17·9%-20·7%]), adults with poor health status (15·6% [95% CI, 11·6%-19·6%]), adults with three or more chronic conditions (24·2% [95% CI, 22·2%-26·2%]), Medicare beneficiaries (22·8% [95% CI, 21·4%-24·3%]), and military-insured adults (13·9% [95% CI, 10·8%-17·1%]). Mail-order prescriptions were commonly filled for analgesics, levothyroxine, cardiovascular agents, antibiotics, and diabetes medications.

**Interpretation:** The use of mail-order prescription, including for critical medications such as insulin, is increasingly common among U.S. adults and displays substantial variation between population groups. A national slowdown of mail delivery could have important health consequences for a considerable proportion of the U.S. population, particularly during the current Coronavirus disease 2019 epidemic.

**Research in context:** *Evidence before this study:* In July 2020, major cost-cutting actions at the United States Postal Service (USPS) caused a sudden slowdown in mail delivery on a national scale. In addition to jeopardizing mail-in ballots for the upcoming November’s general election, such a remarkable postal delay may also deteriorate the health of many individuals who rely on the postal service to deliver their essential medications. The current SARS-CoV-2 epidemic may further amplify the postal crisis given anecdotal evidence that many patients have switched to mail-order prescriptions to avoid potential exposures to the virus at drugstores. Efforts aimed at addressing the health and economic effects of delayed medication delivery require an understanding of the national prevalence of mail-order prescription and the population groups who will likely be most affected by these postal delays. We searched PubMed for articles published on or before September 14, 2020 that described patterns of mail-order medication use using variations of the search terms “mail-order,” “medication,” and “United States.” We found no empirical evidence on trends in the use of mail-order medications at the national level or how such use varied by population subgroups.

*Added value of this study:* To our knowledge, this study is the first to document time trends in the use of mail-order medications using data from a nationally representative cross-sectional sample of U.S. adults for each year from 1996 to 2018. Given the lack of evidence on the use of mail-order medications, the three key contributions of this study are to (i) demonstrate a relative increase of more than 50% in the prevalence of using mail-order prescription from 1996 to 2018, (ii) highlight population groups heavily relying on mail-order prescriptions and who are, thus, most likely to be affected by a decline in the availability or reliability of mail-order prescriptions from cost-cutting measures at the USPS, and (iii) document medications most commonly delivered by mail so that policy makers and clinicians can design effective interventions to alleviate the consequences of delayed or absent mail-order prescriptions.

*Implications of all the available evidence:* We show that the use of mail-order medications has become more prevalent among U.S. adults over the past two decades, with an increase from 13.5 million adults in 1996 to 25.9 million adults in 2018. Given that USPS delivers 55% of all mail-order medications, a considerable proportion of mail-order pharmacy consumers could experience a delay or non-delivery of their medications. The use of mail-order medications is most common among older adults, disabled and chronically ill persons, and military-insured beneficiaries, who may have limited access to a local drugstore due to their morbidities or disability. In addition, we document a variety of medications frequently delivered by mail – ranging from those for which missing several doses does not result in immediate adverse health consequences (e.g. statins), to those for which missing a dose could be detrimental to the patient’s health (e.g. insulin). Our findings highlight that any disruption in the postal service could lead to important medical complications among a considerable proportion of the vulnerable U.S. population. The potential health consequences of a widespread national slowdown in mail delivery should be considered when weighing options to reverse recent changes at the USPS.

## Introduction

Since 2006, the United States Postal Services (USPS) has faced $83·1 billion in cumulative losses,^1^ mainly due to declining mail volumes and a mandate requiring the agency to prefund its employee benefits.^2^ In May 2020, USPS ordered aggressive cost-cutting measures, including slashing overtime delivery trips, sorting machines, and other expenses required to ensure punctual mail delivery.^3^ The National Association of Letter Carriers recommended that the U.S. Congress should provide at least $25 billion in direct financial aid to help strengthen the USPS and reverse recent changes.^2^ However, the U.S. President has recently threatened to veto any congressional postal bills due to concerns about voter fraud related to mail-in ballots.^4^

Studies have consistently found that mail-order medications are associated with reduced healthcare costs and better health outcomes as a result of improvements in medication adherence.^5–7^ However, in a weekly survey conducted in August 2020, one in four mail-order prescription users reported experiencing a delay or non-delivery of their medications during the preceding week.^8,9^ Given anecdotal reports that many patients have switched to mail-order prescriptions since the start of the SARS-CoV-2 epidemic to prevent potential exposure to infection in stores,^10^ demand for mail-order medications – and thus reliance on the USPS mail delivery system – has likely risen over the past months. Delay of critical prescriptions may force patients to visit their local pharmacy, potentially putting them at risk of contracting the virus. Picking up medications from a local drugstore may also increase patients’ copay because many insurance companies cover a higher proportion of the cost when a medication is delivered by mail.^7^ Moreover, many medications are unavailable at the local drugstore because they are a controlled substance or because they are in shortage due to the SARS-CoV-2 epidemic.^11^ Efforts aimed at addressing the health and economic effects of delayed medication delivery require an understanding of the national prevalence of mail-order prescription and the population groups who will likely be most affected by these postal delays. Several studies have estimated the prevalence of mail-order prescription, but they are either outdated,^12,13^ unrepresentative of the U.S. population,^13–15^ or unable to assess variation in prevalence and time trends by socio-demographic and health characteristics.^14,16^

This study uses nationally representative data to determine: (a) the prevalence of mail-order prescription from 1996 to 2018 among U.S. adults, (b) how the prevalence and time trends vary by socio-demographic characteristics, physical access to a usual place for medical care, and health characteristics, and (c) which medications are most likely to be delivered by mail.

## Methods

### Data source and study population

The Medical Expenditure Panel Survey (MEPS) is a nationally representative survey of the civilian noninstitutionalized U.S. population drawn from a subsample of households that responded to the prior year’s National Health Interview Survey (NHIS).^17^ The NHIS is a complex, multistage probability survey conducted in each state and the District of Columbia. In the first stage, a random sample of primary sampling units (PSUs) was selected with the probability of selection being proportionate to a PSU’s population size as estimated by population projections from the U.S. Census Bureau. The second stage consisted of a probability sample of geographic area segments within each PSU. These segments were subsequently divided into clusters, each of which included approximately four to nine housing units. Racial/ethnic minority households were oversampled at a rate of 2:1. Each year, the MEPS randomly selected three-eighths of the households that responded in the prior year’s NHIS. The MEPS’s response rate, after accounting for non-response in the NHIS, ranged from 78% in 1996 to 46·8% in 2018.^18^ MEPS participants enter the survey each year as members of a unique panel. They are interviewed in five sequential rounds spanning two years. Data are organized as annual files, each of which includes members from two unique panels. Analyses were based on adults aged 18 years and older in all available MEPS surveys (1996 to 2018) (N = 507,629). After excluding 7,412 persons with incomplete data on mail-order prescription, the final sample was 500,217. This study is exempt from Institutional Review Board approval because MEPS data are in the public domain.

### Ascertaining mail-order prescription use

In each survey round, respondents reported the name(s) of all prescribed medication(s) that they filled and refilled (whether through the mail or not) during the reference period. The reference periods vary across households, but they typically cover five months on average. Each refill was treated as a single medication record. From 1996 to 2018, approximately 70-80% of respondents signed the permission forms for MEPS to contact their pharmacies, 77% of which agreed to provide information on respondents’ reported medications.^18,19^ All medications were linked to a proprietary database – Lexicon Plus, Cerner Multum – that contains information on all prescription medications available in the U.S. market. The quality of prescription data has been validated elsewhere.^20^ Participants were asked for the type of pharmacy from which each medication was purchased, with the options included mail-order pharmacies; pharmacies located in another store such as a grocery or department store; pharmacies located in an HMO, clinic, or hospital; drug stores that are not located within another facility; or on-line pharmacies. Mail-order prescriptions were defined as those obtained from either a mail-order pharmacy or online store.

### Ascertaining socio-demographic characteristics, physical access to a pharmacy, and health status

MEPS asked participants about the following socio-demographic characteristics: age (categorized in this analysis as 18-44 years, 45-64 years, and 65 years and older), sex, race/ethnicity (non-Hispanic white, non-Hispanic black, non-Hispanic Asian, non-Hispanic others, or Hispanic), education (less than high school, high school graduate, and college graduate), marital status (married, divorced/widowed/separated, and never-married), household poverty level (less than 200%, 200-399%, and 400% or higher of the federal poverty threshold), region of residence (Northeast, Midwest, South, and West), and health insurance coverage (Medicare beneficiaries, Medicaid or public insurance only, military insurance only, private insurance only, and uninsured). To ascertain physical access to the usual place for medical care, respondents were asked for the amount of time it took them to travel to their usual place of medical care, with the response options being less than 15 minutes, 15-30 minutes, 31-60 minutes, and more than 60 minutes. To ascertain health status, participants were asked to rate their general health, with excellent, very good, good, fair, or poor being the response options. In addition, participants were asked whether pain interfered with their normal work last month (none or a bit of the time, and some, most, or all of the time). Limitations in daily activities or routine functions (“any limitation” hereafter) were defined as whether a participant reported any instrumental activities of daily living (IADL) limitation, activities of daily living (ADL) limitation, functional limitation, activity limitation, hearing impairment, or vision impairment. The number of chronic conditions (none, one, two, and three or more) was based on self-report diagnoses of medical conditions. Each condition was recorded as verbatim text then coded to ICD-9 codes. We defined the following conditions as chronic: hypertension, congestive heart failure, coronary artery disease, cardiac arrhythmias, hyperlipidemia, stroke, arthritis, asthma, cancer, chronic kidney disease, chronic obstructive pulmonary disease, dementia, diabetes, hepatitis, human immunodeficiency virus (HIV), and osteoporosis.^21^

### Statistical analysis

We estimated the prevalence of using any mail-order prescription and the mean number of mail-order prescriptions in a calendar year using sampling weights that accounted for the survey design. The variance of these estimates was calculated using the Taylor series linearization method. Both estimates were age-standardized to the population age distribution of the U.S. in 2000 as published by the U.S. National Census Bureau (Appendix Figure 1).^22^ All estimates were stratified by socio-demographic characteristics, physical access to the usual place for medical care, and health status. We reported the prevalence of mail-order prescription use among the general adult population for the 25 medications that constituted the largest proportion of all medications delivered by mail. In addition, we reported the prevalence of use for the 25 medications that were most commonly delivered by mail among the users of these medications, conditional on having at least 500 users in the pooled 1996-2018 data to allow for reasonably precise estimates.

**Figure 1:**
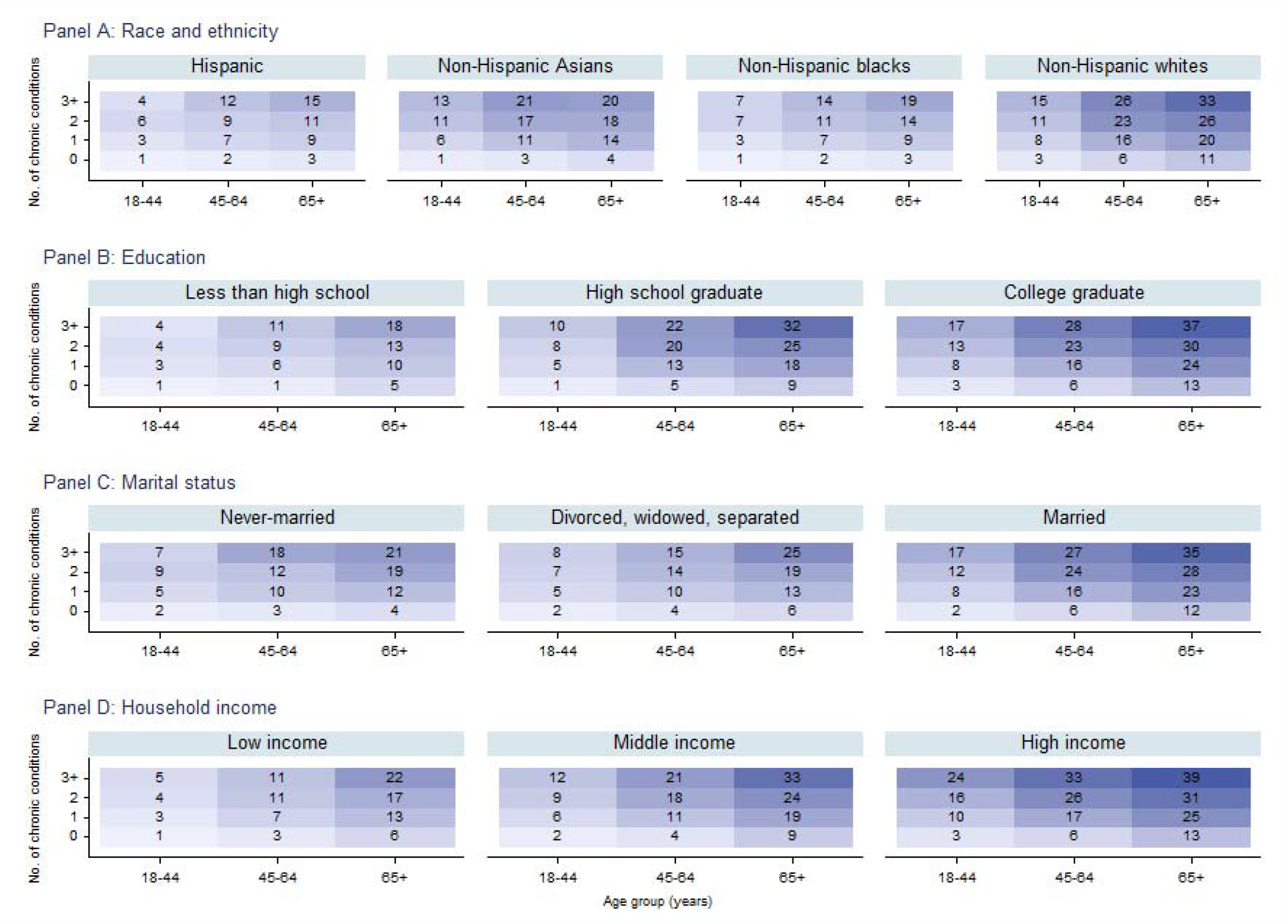
Prevalence of mail-order prescription use among U.S. adults by age group, number of chronic conditions, and other selected socio-demographic characteristics – 1996-2018. Note: Prevalence of mail-order prescription use is presented in each cell.

All analyses were conducted using Stata version 13.0.

## Results

### Sample characteristics and prevalence of mail-order prescription use

Table 1 presents descriptive statistics for socio-demographic and health characteristics of the full sample and by the use of mail-order prescription. Overall, 9·8% (95% CI, 9·5%-10·0%) of adults used at least one mail-order prescription in a year. Among these individuals, each user filled a mean of 19·4 (95% CI, 19·2-19·6) mail-order prescriptions per year. Compared to nonusers, mail-order prescription users were more likely to be older, female, non-Hispanic white, college educated, married, high-income, reporting poor or fair health, experiencing pain interference with their normal work, having any limitation, reporting multiple chronic conditions, Medicare beneficiaries, and having a usual place for medical care.

**Table 1:** Prevalence of mail-order prescription use among U.S. adults – 1996-2018.

|  | (1)<br>Total sample | (2)<br>No mail-order<br>prescription use | (3)<br>Any mail-order<br>prescription use |
| --- | --- | --- | --- |
|  | No. of<br>participants (%<sup>a,b</sup>)<br>N = 500217 | No. (prevalence in %<sup>a,b</sup>) [95% CI] who used mail-order<br>prescription in a year<br>N = 460086 | N = 40131 |
| **Sample size** |  |  |  |
| Used any mail-order prescription | 40131 (9.8) | 0 (0) | 40131 (100) |
| No. of mail-order<br>prescriptions, mean<sup>a,c</sup> [95% CI] | 1.9 [1.8, 2.0] | 0 [0, 0] | 19.4 [19.0, 19.8] |
| **Socio-demographic variables** |  |  |  |
| Age groups |  |  |  |
| 18-44 | 251902 (49.3) | 245717 (52.8) [52.4, 53.3] | 6185 (16.4) [15.8, 17.0] |
| 45-64 | 163275 (32.9) | 146528 (32.0) [31.7, 32.3] | 16747 (41.4) [40.5, 42.4] |
| 65+ | 85040 (17.8) | 67841 (15.2) [14.9, 15.5] | 17199 (42.2) [41.1, 43.2] |
| Sex |  |  |  |
| Men | 230339 (48.1) | 213133 (48.6) [48.4, 48.8] | 17206 (43.6) [43.0, 44.2] |
| Women | 269878 (51.9) | 246953 (51.4) [51.2, 51.6] | 22925 (56.4) [55.8, 57.0] |
| Race/ethnicity |  |  |  |
| Non-Hispanic white | 257530 (68.4) | 226890 (66.6) [65.5, 67.6] | 30640 (85.8) [85.0, 86.5] |
| Non-Hispanic black | 82928 (11.5) | 78795 (12.1) [11.4, 12.8] | 4133 (5.7) [5.2, 6.1] |
| Non-Hispanic Asian | 28798 (4.9) | 27147 (5.1) [4.7, 5.5] | 1651 (2.9) [2.5, 3.3] |
| Non-Hispanic others | 8816 (1.8) | 8236 (1.8) [1.6, 2.0] | 580 (1.3) [1.1, 1.5] |
| Hispanic | 122145 (13.5) | 119018 (14.4) [13.6, 15.3] | 3127 (4.4) [4.0, 4.8] |
| Educational attainment<sup>d</sup> |  |  |  |
| Less than high school | 115209 (16.2) | 110589 (17.0) [16.6, 17.4] | 4620 (9.0) [8.5, 9.4] |
| High school graduate | 161754 (32.3) | 148410 (32.3) [31.9, 32.7] | 13344 (32.0) [31.1, 32.9] |
| College or above | 219498 (51.5) | 197462 (50.7) [50.1, 51.3] | 22036 (59.0) [57.9, 60.1] |
| Marital status<sup>e</sup> |  |  |  |
| Married | 263451 (54.2) | 236815 (52.8) [52.3, 53.2] | 26636 (67.5) [66.6, 68.4] |
| Divorced, widowed, or separated | 102704 (19.9) | 92864 (19.5) [19.2, 19.8] | 9840 (23.1) [22.4, 23.9] |
| Never-married | 134047 (25.9) | 130392 (27.7) [27.4, 28.1] | 3655 (9.3) [8.8, 9.8] |
| Household poverty level<sup>f</sup> |  |  |  |
| Low-income | 179268 (28.0) | 170882 (29.1) [28.6, 29.7] | 8386 (18.2) [17.6, 18.9] |
| Middle-income | 145067 (30.4) | 134140 (30.7) [30.4, 31.1] | 10927 (27.6) [26.8, 28.4] |
| High-income | 153863 (41.5) | 135301 (40.1) [39.5, 40.8] | 18562 (54.2) [53.1, 55.3] |
| Region of residence<sup>g</sup> |  |  |  |
| Northeast | 81414 (18.6) | 74055 (18.4) [17.3, 19.4] | 7359 (20.6) [18.9, 22.2] |
| Midwest | 99250 (22.0) | 89498 (21.8) [20.8, 22.8] | 9752 (24.0) [22.4, 25.6] |
| South | 188945 (36.4) | 174803 (36.6) [35.4, 37.8] | 14142 (34.9) [33.2, 36.7] |
| West | 130604 (23.0) | 121726 (23.2) [22.2, 24.3] | 8878 (20.5) [19.2, 21.9] |
| **Health characteristics and access to care** |  |  |  |
| Self-reported health status<sup>h</sup> |  |  |  |
| Excellent | 70152 (19.0) | 66959 (20.1) [19.7, 20.4] | 3193 (10.3) [9.8, 10.8] |
| Very good | 130808 (36.5) | 119498 (36.7) [36.3, 37.0] | 11310 (35.2) [34.4, 36.0] |
| Good | 124694 (30.5) | 112131 (29.8) [29.5, 30.1] | 12563 (36.4) [35.7, 37.1] |
| Fair | 52871 (11.3) | 47263 (10.9) [10.7, 11.1] | 5608 (14.6) [14.0, 15.2] |
| Poor | 12799 (2.7) | 11363 (2.6) [2.5, 2.7] | 1436 (3.4) [3.2, 3.7] |
| Frequency pain interfered with work<sup>i</sup> |  |  |  |
| None or a bit of the time | 306581 (79.5) | 282855 (80.5) [80.2, 80.8] | 23726 (71.4) [70.7, 72.2] |
| Some, most, or all of the time | 85008 (20.5) | 74563 (19.5) [19.2, 19.8] | 10445 (28.6) [27.8, 29.3] |
| Any limitation<sup>j</sup> |  |  |  |
| No | 368106 (74.6) | 345252 (76.3) [75.9, 76.7] | 22854 (58.8) [57.9, 59.6] |
| Yes | 126640 (25.4) | 109519 (23.7) [23.3, 24.1] | 17121 (41.2) [40.4, 42.1] |
| Number of chronic conditions<sup>k</sup> |  |  |  |
| None | 245346 (55.5) | 239485 (59.5) [59.1, 60.0] | 5861 (17.9) [17.2, 18.6] |
| One | 86035 (20.8) | 77463 (20.3) [20.1, 20.5] | 8572 (25.3) [24.6, 26.0] |
| Two | 46574 (11.1) | 38732 (9.9) [9.7, 10.0] | 7842 (22.7) [22.1, 23.3] |
| Three or more | 53577 (12.6) | 42016 (10.3) [10.0, 10.6] | 11561 (34.1) [33.2, 35.0] |
| Health insurance |  |  |  |
| Medicare | 97117 (19.7) | 78837 (17.1) [16.7, 17.4] | 18280 (44.0) [43.0, 45.1] |
| Medicaid or public insurance only | 62069 (9.0) | 60686 (9.7) [9.4, 10.0] | 1383 (3.0) [2.7, 3.2] |
| Private insurance only | 248103 (56.8) | 229431 (57.7) [57.1, 58.3] | 18672 (49.2) [48.1, 50.2] |
| Military insurance only | 7591 (1.6) | 6853 (1.6) [1.5, 1.7] | 738 (1.7) [1.5, 2.0] |
| Not insured | 85337 (12.8) | 84279 (13.9) [13.5, 14.3] | 1058 (2.1) [1.9, 2.3] |
| Travel time to usual place of medical care<sup>l</sup> |  |  |  |
| No usual source of medical care | 97022 (23.4) | 95292 (25.6) [25.0, 26.1] | 1730 (5.6) [5.2, 6.0] |
| < 15 mins | 131966 (38.7) | 118158 (37.9) [37.3, 38.6] | 13808 (45.0) [43.7, 46.4] |
| 15-30 mins | 105516 (30.1) | 93382 (29.0) [28.5, 29.6] | 12134 (39.0) [37.8, 40.2] |
| 31-60 mins | 24116 (6.5) | 21295 (6.3) [6.0, 6.5] | 2821 (8.6) [8.1, 9.2] |
| > 60 mins | 4832 (1.3) | 4245 (1.2) [1.1, 1.3] | 587 (1.7) [1.4, 1.9] |

a: Weighted estimates using sampling weight.
b: Estimates may not add up to 100% due to rounding error.
c: Based on 40,131 respondents who reported using at least one mail-order prescription in a calendar year.
d: Excludes 3756 respondents whose educational attainment was not collected.
e: Excludes 15 respondents whose marital status was not collected.
f: Households were classified into three categories based on their total income and household composition: low-income (less than 200% of the federal poverty threshold), middle-income (200-399%), and high-income ( ≥ 400%).
g: Excludes 4 respondents whose region of residence was not collected.
h: Excludes 108893 respondents whose health status was not collected. MEPS started collecting this variable in 2000. Respondents interviewed before 2000 were removed for this particular variable.
i: Excludes 108628 respondents whose information on pain interference was not collected. MEPS started collecting this variable in 2000. Respondents interviewed before 2000 were removed for this particular variable.

The prevalence of mail-order prescription use varied markedly between population groups (Figure 1). Among adults aged 65 and older who reported having three or more chronic conditions, 33% of non-Hispanic whites (Panel A), 37% of college graduates (Panel B), 35% of married adults (Panel C), and 39% of high-income adults (Panel D) used mail-order prescription. More generally, the prevalence of mail-order prescription use was particularly high among older adults (23·1% [95% CI, 22·5%-23·8%]), non-Hispanic whites (12·2% [95% CI, 11·9%-12·6%]), college graduates (11·2% [95% CI, 10·9%-11·6%]), married adults (12·2% [95% CI, 11·8%-12·5%]), divorced, widowed, or separated adults (11·4% [95% CI, 11·0%-11·8%]), high-income adults (12·7% [95% CI, 12·3%-13·1%]), adults reporting fair (14·0% [95% CI, 13·4%-14·6%]) or poor (13·7% [95% CI, 12·8%-14·8%]) health, adults with any limitation (15·9% [95% CI, 15·5%-16·4%]), adults with three or more chronic conditions (26·1% [95%, 25·3%-26·9%]), Medicare beneficiaries (21·8% [95% CI, 21·3%-22·4%]), and adults whose travel time to their usual place for medical care was more than 60 minutes (14·7% [95% CI, 13·1%-16·3%]) (Appendix Table 1).

### Time trends in mail-order prescription use

The prevalence of mail-order prescription use increased from 6·9% (95% CI, 6·4%-7·5%) in 1996 to 11·4% (95% CI, 10·6%-12·1%) in 2005 (Appendix Figure 1, Panel A), stagnated between 2006 and 2011, then declined after 2011, with the prevalence in 2018 being 10·3% (95% CI, 9·7%-10·9%). Each user purchased a mean of 10·7 (95% CI, 9·8-11·7) mail-order prescriptions in 1996, which increased to 20·5 (95% CI, 19·3-21·7) in 2018 (Appendix Figure 1, Panel B). Age-standardized estimates were lower than crude estimates, although the patterns for prevalence of use and for the number of mail-order prescriptions remained unchanged.

In Figure 2, gender differences in mail-order prescription use were small throughout 1996 to 2018 (Panel A). The prevalence of mail-order prescription use was low among young adults (aged 18 to 44 years) for all years from 1996 to 2018, while it increased between 1996 and 2006 among adults aged 45 years and older to then gradually decline from 2006 onwards (Panel B). Among adults aged 65 years and older, the prevalence of mail-order prescription use peaked in 2010 at 28·9% (95% CI, 25·8%-30·0%) and has since decreased to 23·9% (95% CI, 22·3%-25·4%) in 2018. By race/ethnicity, mail-order prescription use was most prevalent among non-Hispanic whites and least prevalent among Hispanics from 1996 to 2018 (Panel C).

**Figure 2:**
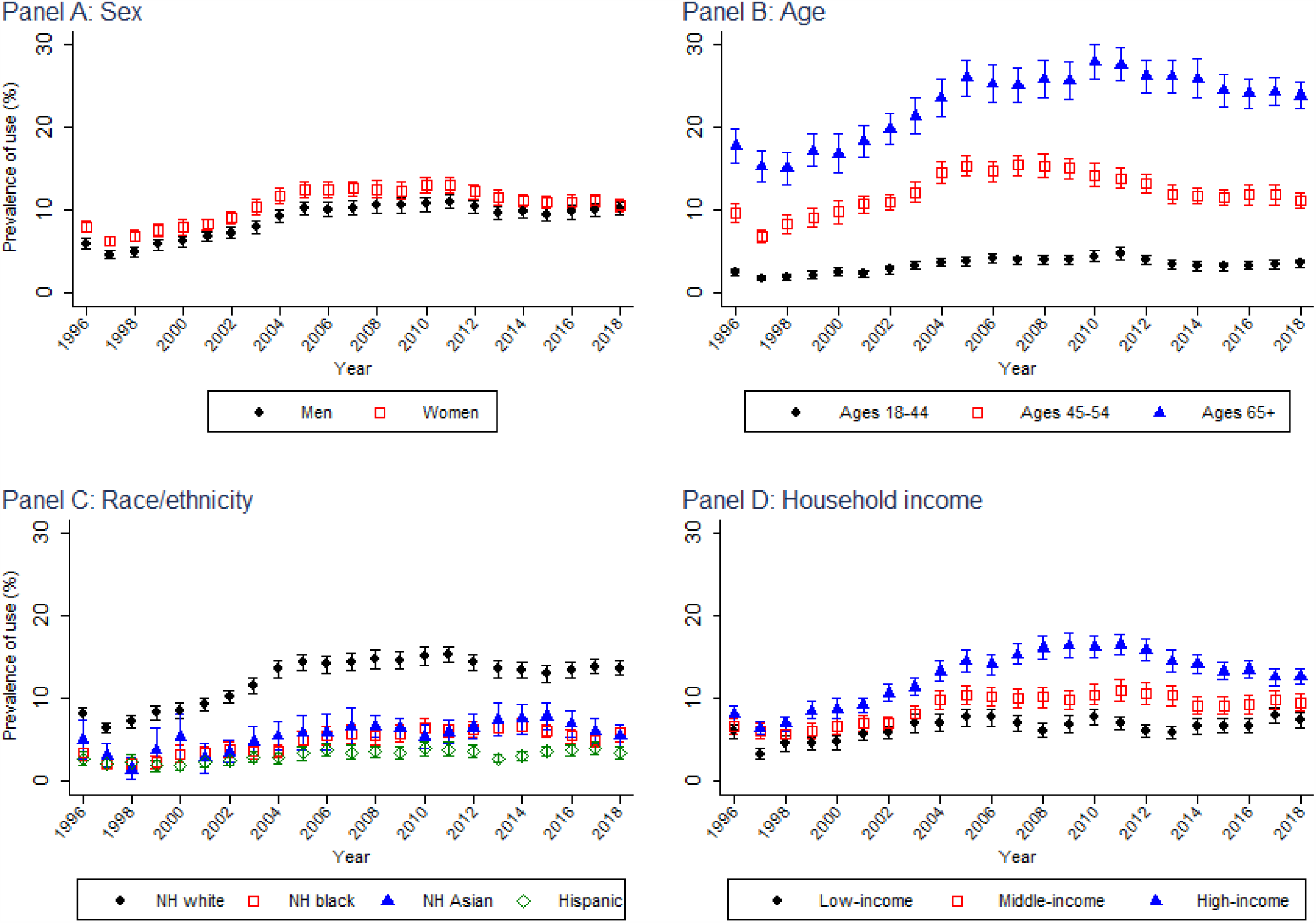
Trends in mail-order prescription use among U.S. adults by sex, age, race/ethnicity, and household income – 1996-2018. Abbreviation: NH, non-Hispanic Note: All estimates were not age-standardized.

When stratifying prevalence of mail-order prescription use by household income, prevalence was higher among high-income than middle- and low-income adults for all years from 1996 to 2017 (Figure 2, Panel D). However, the use of mail-order prescription generally increased between 1996 and 2018 among low-(6·0% in 1996, 7·3% in 2018, difference of 1·4 percentage points [95% CI, 0·1-2·6 percentage points]) and middle-income (6·6% in 1996, 9·5% in 2018, difference of 2·9 percentage points [95% CI, 1·5-4·2 percentage points]) adults, whereas it decreased among high-income adults from 16·5% in 2011 to 12·6% in 2018 (difference of −3·9 percentage points [95% CI, −5·5 to −2·3 percentage points]). In 2018, differences in the prevalence of mail-order prescription use between income groups were comparatively small among high-income (12·6 % [95% CI, 11·6%-13·6%]), middle-income (9·5% [95% CI, 8·5%-10·5%]), and low-income adults (7·3% [95% CI, 6·4%-8·2%]).

The prevalence of mail-order prescription use among other subpopulations generally increased during the first ten years of the study period and then declined. In Appendix Figure 2, the prevalence of use among married, divorced, widowed, or separated adults (Panel A), high school and college graduates (Panel B); adults residing in each of the four regions of the U.S. (Panel C) generally increased during the 1996-2006 period, stagnated from 2006 to 2010, and then declined from 2010 onwards. We observed a similar pattern in Appendix Figure 3 for adults of any health status (Panel C); adults with one or more chronic conditions (Panel D); and Medicare beneficiaries and privately insured adults (Panel E). Population groups with a consistently low prevalence of mail-order prescription use throughout the study period included adults with less than a high school education (Appendix Figure 2); as well as adults without chronic conditions, Medicaid or publicly insured adults, uninsured adults, and adults without a usual place for medical care (Appendix Figure 3). Time trends in the prevalence of mail-order prescription use by limitation and pain interference are shown in Appendix Figure 3, and by type of limitation in Appendix Figure 4.

### Mail-order prescription use by medication

Among the general U.S. adult population, mail-order prescriptions were commonly filled for analgesics (e.g. acetaminophen-hydrocodone [6·3%] and ibuprofen [4·0%]), thyroid hormones (e.g. levothyroxine [6·1%]), cardiovascular agents (e.g. lisinopril [5·8%], metoprolol [4·3%], and amlodipine [3·6%]), anti-infective agents (e.g. azithromycin [5·6%] and amoxicillin [5·1%]), antihyperlipidemic agents (e.g. atorvastatin [5·2%] and simvastatin [4·7%]), and anti-diabetic agents (e.g. metformin [4·2%]) (Table 2 and Appendix Table 2). Medications most commonly purchased by mail among their users are listed in Table 3 and Appendix Table 3. Adults who use antihyperlipidemic medications were most likely to obtain these by a mail-order prescription, with between 32·5% to 35·9% of those using ezetimibe, ezetimibe-simvastatin, and niacin receiving these medications in the mail. Other commonly mailed medications – all of which have a prevalence of mail-order prescription use of about one-third among their users – included estrogen, cardiovascular agents (e.g. hydrochlorothiazide-irbesartan, nadolol, and irbesartan), bone resorption inhibitors (e.g. ibandronate, alendronate, and risedronate), and topical estradiol.

**Table 2:** Prevalence of mail-order prescription use among U.S. adults for the 25 most commonly mailed medications – 1996-2018.

|  | Percent of all adults who received a mail-order<br>prescription for the medication (95% CI)<sup>a</sup> |
| --- | --- |
|  | N = 499372<sup>b</sup> |
| Acetaminophen-Hydrocodone | 6.3 (6.1, 6.5) |
| Levothyroxine | 6.1 (5.9, 6.2) |
| Lisinopril | 5.8 (5.6, 5.9) |
| Azithromycin | 5.6 (5.5, 5.8) |
| Atorvastatin | 5.2 (5.1, 5.4) |
| Amoxicillin | 5.1 (5.0, 5.2) |
| Simvastatin | 4.7 (4.6, 4.9) |
| Metoprolol | 4.3 (4.2, 4.5) |
| Metformin | 4.2 (4.1, 4.3) |
| Ibuprofen | 4.0 (3.9, 4.1) |
| Albuterol | 3.8 (3.7, 3.9) |
| Amlodipine | 3.6 (3.5, 3.7) |
| Omeprazole | 3.6 (3.5, 3.7) |
| Hydrochlorothiazide | 3.6 (3.5, 3.7) |
| Prednisone | 3.2 (3.1, 3.2) |
| Atenolol | 2.8 (2.7, 2.9) |
| Furosemide | 2.7 (2.7, 2.8) |
| Naproxen | 2.3 (2.2, 2.4) |
| Acetaminophen-Oxycodone | 2.2 (2.1, 2.3) |
| Cephalexin | 2.2 (2.1, 2.2) |
| Cyclobenzaprine | 2.2 (2.1, 2.2) |
| Sertraline | 2.2 (2.1, 2.2) |
| Potassium Chloride | 2.1 (2.1, 2.2) |
| Acetaminophen-Codeine | 2.0 (2.0, 2.1) |
| Fluticasone Nasal | 2.0 (1.9, 2.1) |

a: The numerator was the number of adult MEPS participants who obtained a mail-order prescription for the medication, and the denominator was all adult MEPS participants.
b: Excluded 845 respondents from the analytic sample of 500,217 persons who had missing information on medication name(s).
Abbreviation: CI, confidence interval.

**Table 3:** Prevalence of mail-order prescription use among adults who have been prescribed each medication, for the 25 medications with the highest prevalence – 1996-2018.

|  | No. of users | Percent of users of the medication who obtained a mail-order prescription (95% CI)<sup>a</sup> |
| --- | --- | --- |
| Ezetimibe | 2420 | 35.9 (33.0, 38.8) |
| Ezetimibe-Simvastatin | 2001 | 34.7 (31.5, 38.0) |
| Raloxifene | 1439 | 34.3 (30.9, 37.6) |
| Hydrochlorothiazide-Irbesartan | 680 | 34.2 (30.5, 37.9) |
| Ibandronate | 615 | 32.6 (29.2, 36.1) |
| Niacin | 1616 | 32.5 (29.6, 35.3) |
| Dutasteride | 552 | 32.3 (28.6, 35.9) |
| Calcitonin | 534 | 31.9 (27.3, 36.4) |
| Alendronate | 5400 | 31.3 (29.4, 33.2) |
| Risedronate | 1403 | 31.1 (28.2, 34.0) |
| Cyclosporine Ophthalmic | 783 | 30.8 (27.0, 34.6) |
| Calcitriol | 545 | 30.8 (26.6, 35.0) |
| Estradiol Topical | 951 | 30.7 (27.3, 34.2) |
| Nadolol | 562 | 30.6 (27.4, 33.8) |
| Irbesartan | 1646 | 30.0 (26.9, 33.2) |
| Torsemide | 730 | 29.8 (26.3, 33.4) |
| Pioglitazone | 3405 | 29.8 (26.9, 32.7) |
| Rosuvastatin | 5343 | 29.8 (27.8, 31.8) |
| Desloratadine | 1045 | 29.6 (26.0, 33.3) |
| Omega-3 Polyunsaturated Fatty Acids | 1175 | 29.6 (26.6, 32.7) |
| Fenofibrate | 3506 | 29.5 (27.1, 31.9) |
| Mesalamine | 914 | 29.3 (25.6, 33.0) |
| Esomeprazole | 6500 | 29.3 (27.3, 31.3) |
| Ramipril | 2537 | 29.3 (26.5, 32.0) |
| Azelastine Nasal | 1093 | 29.2 (25.7, 32.7) |

a: The numerator was the number of adult MEPS participants who obtained a mail-order prescription for the medication, and the denominator was the number of adult MEPS participants who have been prescribed the medication.
Abbreviation: CI, confidence interval.

## Discussion

From 1996 to 2018, the use of any mail-order prescription grew from 6·9% to 10·3%, an increase of 48·6% relative to the baseline. Among users, the number of mail-order medications increased by almost two-fold between 1996 and 2018. Mail-order prescriptions were most commonly used among older adults, adults of high socioeconomic status, disabled and chronically ill persons, and military-insured beneficiaries. While mail-order prescription use was more common among high-income adults than middle- and low-income adults, the differences in mail-order prescription use between income groups have narrowed in recent years. Medications most commonly delivered by mail included those for which missing several doses does not result in immediate adverse health consequences (e.g. statins), but also those for which missing a dose could be detrimental to the patient’s health (e.g. insulin), produce considerable discomfort (e.g. analgesics), have negative consequences for public health (e.g. antibiotics for which delayed or lacking mail-order prescriptions may result in incomplete antibiotic courses and, thus, an increased risk of antibiotic resistance), or that could be critical in sudden deteriorations of a chronic condition (e.g. albuterol).

To our knowledge, this study is the first to use a nationally representative survey of U.S. adults to assess the prevalence of mail-order prescription over the past two decades. Ma and Wang have previously used MEPS to estimate the prevalence of mail-order prescription use but they only used the 2012 wave of the survey and excluded respondents who entirely filled their prescriptions at Health Maintenance Organizations, clinics, or hospitals throughout the year.^12^ Other studies have used non-representative samples, such as online surveys or studies that only sampled a particular population subgroup.^13,15,23^

Our study found that the use of mail-order prescription has slowed down among the adult population, despite the rapid increase prior to 2006. From 1996 to 2005, the prevalence of use increased from 6·9% to 11·4%. It stagnated from 2006 to 2011 at around 11-12%, then declined to 10·3% in 2018. The implementation of Medicare Part D in 2006 that prohibited the mandatory use of mail-order pharmacies may partially be responsible for this pattern.^24^ Particularly, Part D plans that offer their beneficiaries prescription benefits at mail-order pharmacies must provide the same benefits at retail pharmacies.

Given 83·7% of older adults opposing mail-order pharmacies, this requirement likely is an important reason for which many Medicare beneficiaries opted against mail-order prescription benefits.^23^ In addition, and unrelated to the implementation of Medicare Part D, fewer employers have required mail-order services for filling 90-day prescriptions for maintenance medications, with a decline from 45% in 2011 to 23% in 2012.^25^ This shift likely reduced the cost gap between mail and retail pharmacies, which might partially explain why fewer privately-insured adults purchased mail-order prescriptions.

This study identified several subpopulations that rely heavily on mail-order prescriptions. It is likely that a deterioration in the availability or reliability of mail-order prescriptions from cost-cutting measures at the USPS will affect these population groups the most. In particular, adults in the poorest health will likely be hit hardest by USPS cost-cutting measures because they are physically least able to travel to the pharmacy to obtain their medications and our analysis showed them to be most probable to be using mail-order prescriptions. Importantly, it is also these adults who will be at the highest risk of a fatal disease course if they acquire a SARS-CoV-2 infection while in, or traveling to, the pharmacy.^26^ Given that we found that a wide variety of prescribed medications are commonly delivered through the mail, the immediate consequences of delayed or absent mail-order prescriptions could range from poorer adherence to preventive medications to an increased incidence of health emergencies, such as diabetic ketoacidosis or acute exacerbations of asthma.

The social equity implications of reduced availability and reliability of mail-order prescriptions are likely to be profound. While we found that high-income adults still use mail-order prescription medications more heavily than middle- and low-income adults, the differences in use between income groups have narrowed considerably over recent years. Indeed, a simple linear extrapolation of the trends since 2014 suggests that both middle- and low-income adults may already be having a higher prevalence of mail-order prescription use in 2020 than high-income adults. Regardless, it is likely that, on average, individuals with lower income and less education will find it more difficult to change from receiving medications through the USPS to receiving them through another postal carrier or picking up medications in person. The potential reasons are numerous, including a lower ability to bear additional costs from such a change, less flexible employment situations to allow for time to pick up medications, and lower health literacy.^7,27^

While our study is unique in its use of a nationally representative repeated cross-sectional sample of adults for each year spanning more than two decades, it faces some limitations. First, we were not able to determine the prevalence of using mail-order pharmacy for over-the-counter medications because MEPS only collected data on prescription medications. Second, our measure of using mail-order prescriptions was based on self-report and may, thus, be affected by recall bias. However, among those who reported using any prescriptions, the MEPS team was able to contact the pharmacy of over half of participants to verify their answers. Third, while the response rate of the NHIS and MEPS is high compared to other health surveys in the U.S.,^28,29^ it is possible that adults who were selected for the survey but were not reached or declined to participate had a systematically different prevalence of mail-order prescription use than MEPS participants. The sampling weights in the MEPS accounted for non-response using participants’ measured socio-demographic characteristics. Finally, we were unable to examine the prevalence of mail-order prescription delivered by USPS because MEPS did not collect data on the service carrier. It has been estimated that USPS delivers 55 percent of the nation’s medications.^30^

## Conclusion

The use of mail-order prescription is common among U.S. adults, particularly among those in poorer health, and has increased substantially since 1996. Our analysis highlights that any disruption in the postal service could lead to important medical complications among a considerable proportion of the U.S. population. These health consequences should be taken into account when weighing operational and funding changes to the USPS.

## Data Availability

Data are available at https://www.meps.ahrq.gov/mepsweb/

## Contributors

DD and PG conceived and designed the study. DD processed the data, conducted the analyses, and visualized the data. DD and PG interpreted the results. Both authors wrote the first draft of the manuscript, critically revised the article, and approved the final version.

## Acknowledgement

PG was supported by the National Center for Advancing Translational Sciences of the National Institutes of Health under Award Number KL2TR003143.

---

**Appendix Figure 1:**
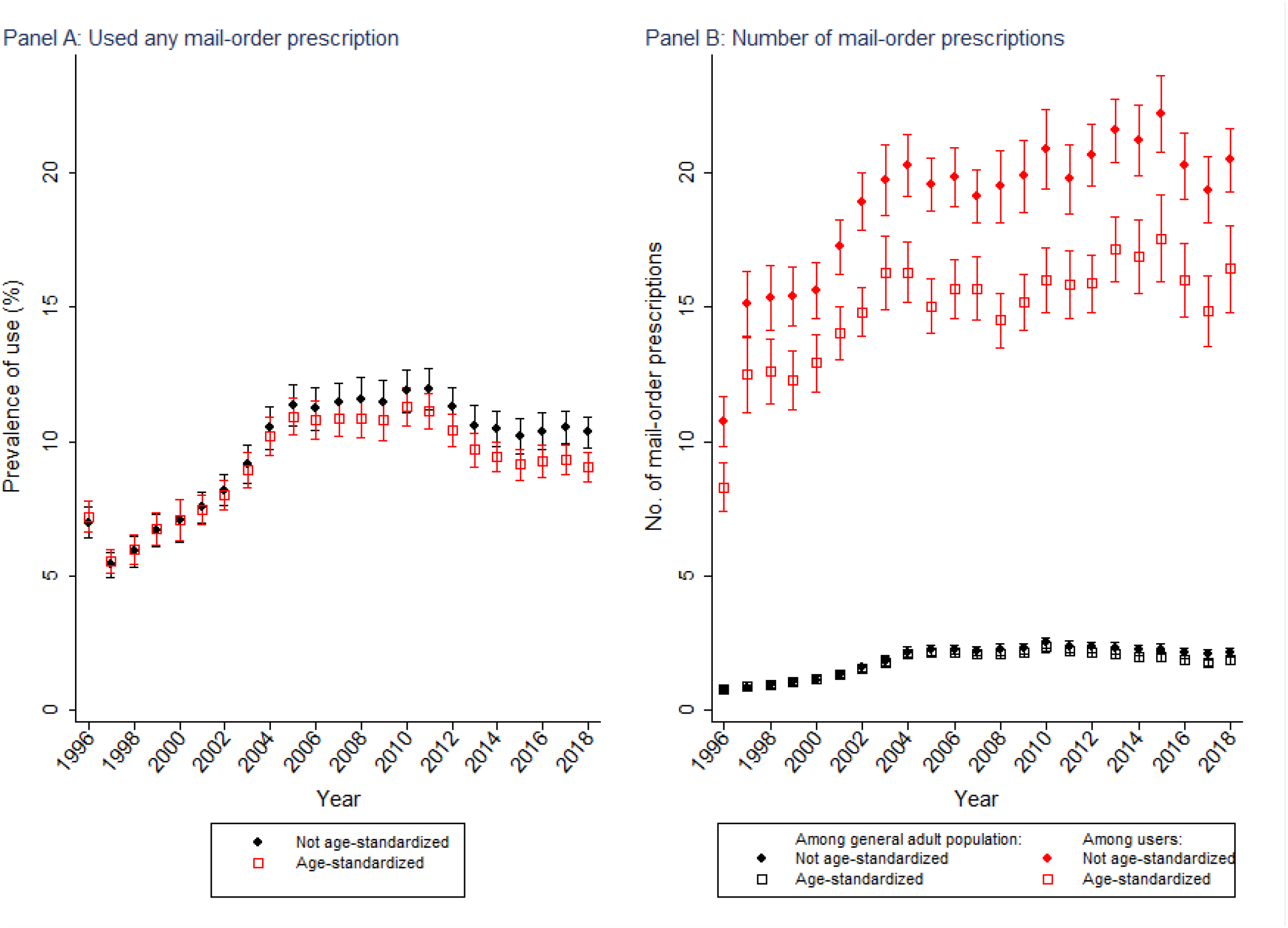
Trends in mail-order prescription use and the number of mail-order prescriptions among U.S. adults – 1996-2018.

**Appendix Figure 2:**
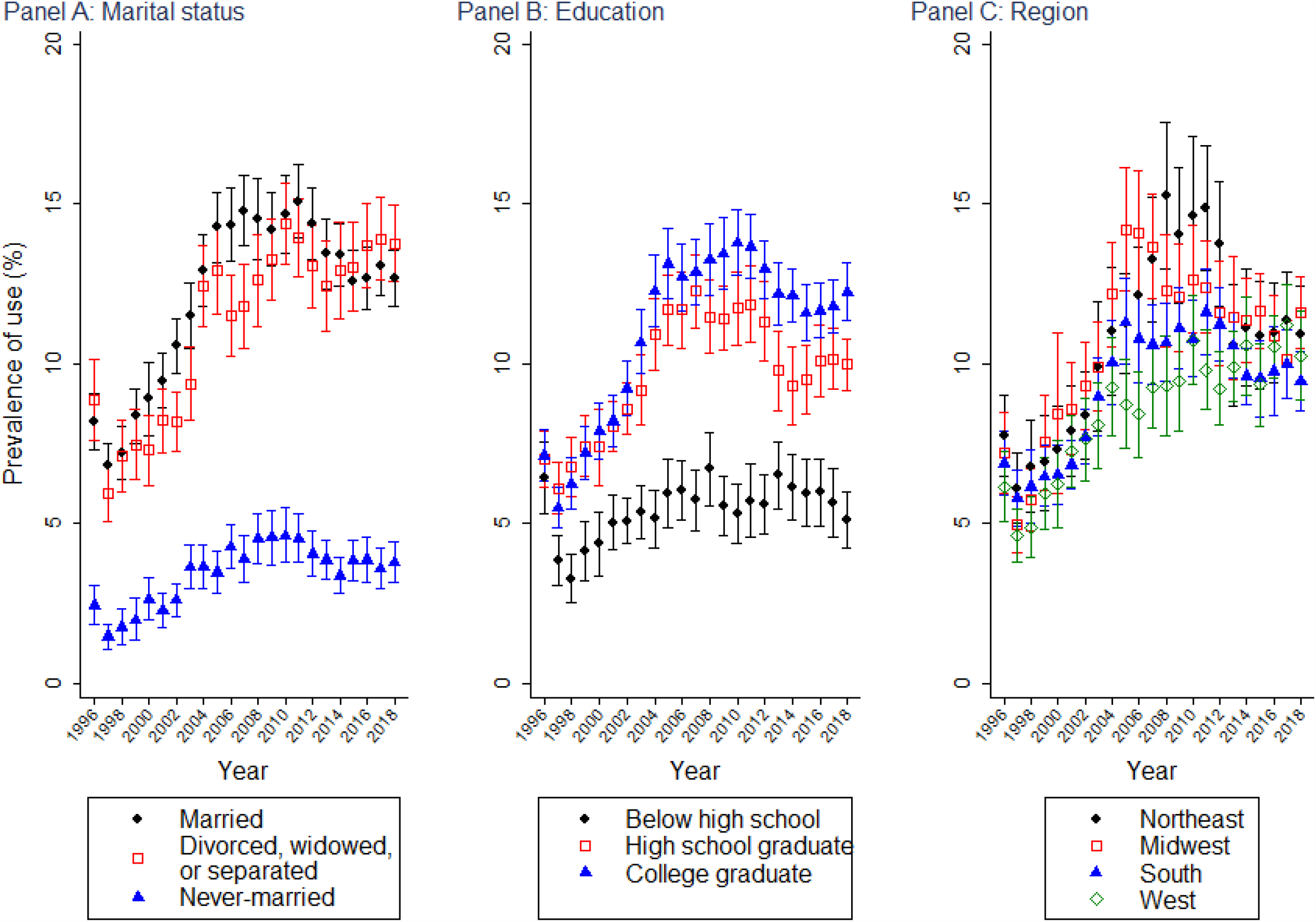
Trends in mail-order prescription use by marital status, education, and region – 1996-2018. Note: All estimates were not age-standardized.

**Appendix Figure 3:**
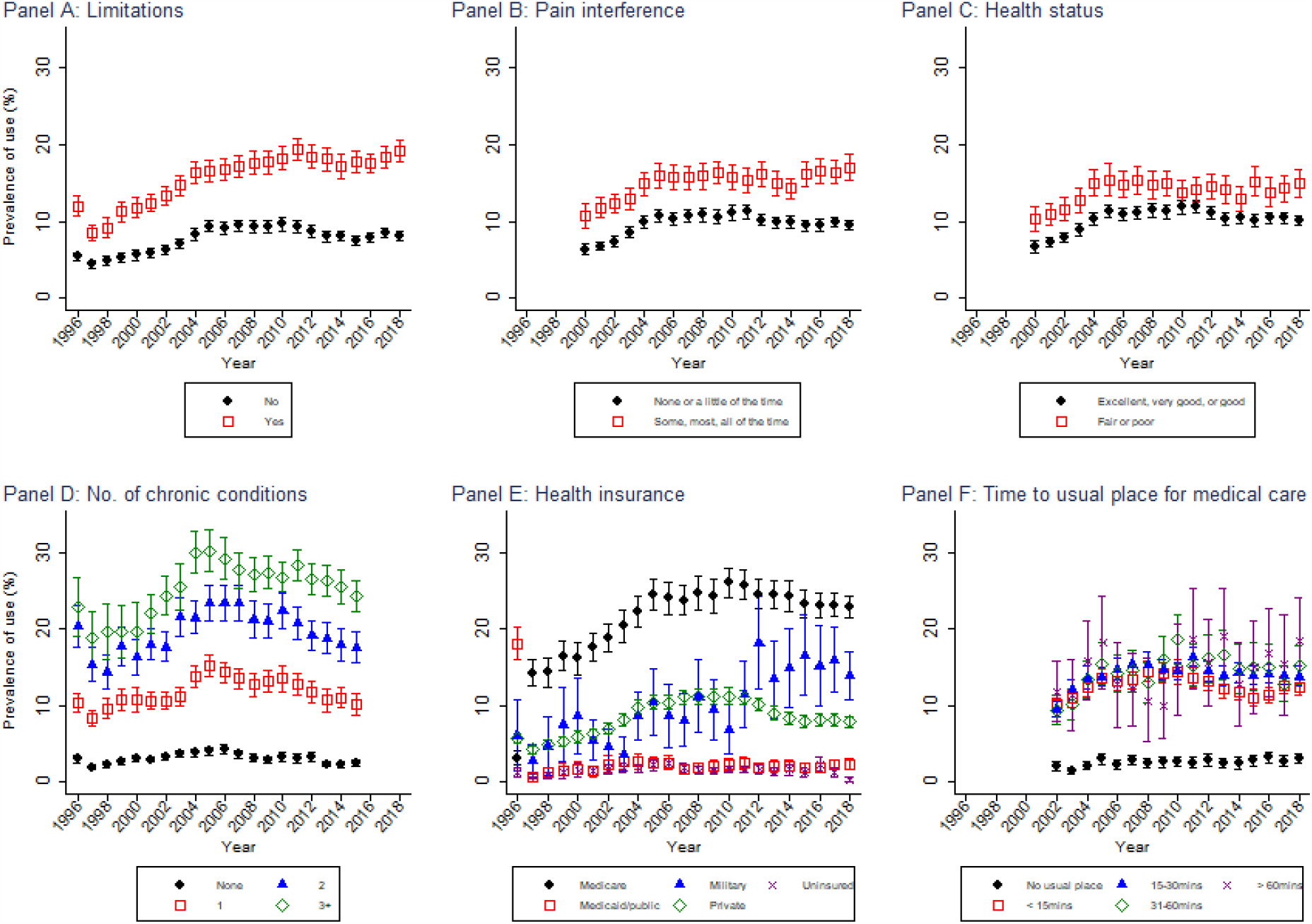
Trends in mail-order prescription use among U.S. adults by health characteristics and access to care – 1996-2018. Note: All estimates were not age-standardized. Panel B: Excludes 108628 respondents whose information on pain interference was not collected. MEPS started collecting this variable in 2000. Panel C: Excludes 108893 respondents in 1996-1999 whose health status was not collected. MEPS started collecting this variable in 2000. Panel D: Excludes 68685 respondents in 2016-2018 whose information on chronic conditions was not collected. In 2016, MEPS started using the International Classification of Diseases 10 (ICD-10) instead of ICD-9. As such, the medical classifications of conditions in 2016 are still under review. Panel F: Excludes 114,774 respondents whose travel time to the usual place of medical care was not collected. MEPS started collecting this variable in 2002.

**Appendix Figure 4:**
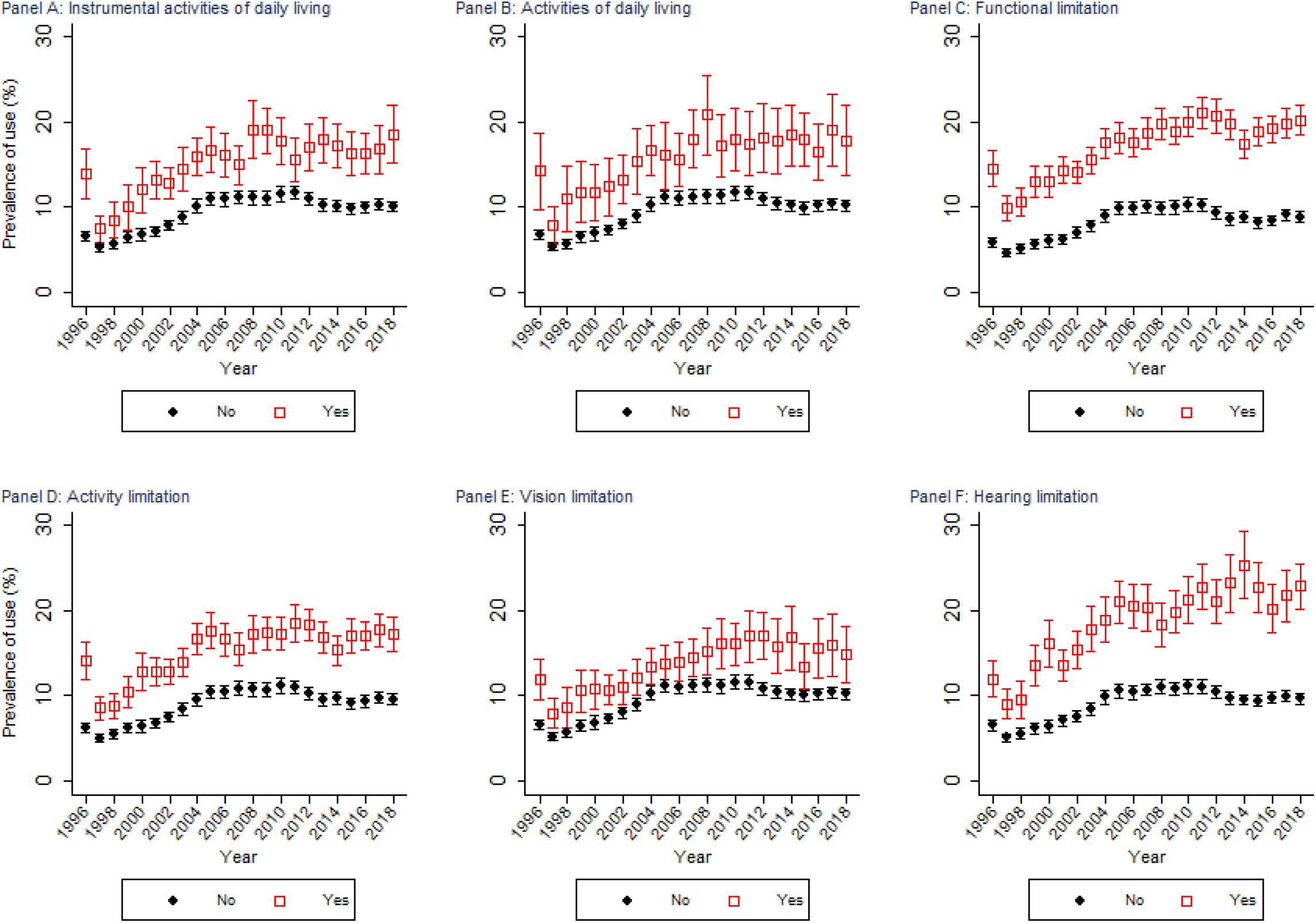
Trends in mail-order prescription use among U.S. adults by type of limitation – 1996-2018.

**Appendix Table 1:** Prevalence of mail-order prescription use among U.S. adults stratified by socio-demographic and health characteristics – 1996-2018.

| Characteristics | (1)<br>No. of<br>participants | (2)<br>Used any mail-order<br>prescriptions<br>Prevalence of use, %<sup>a</sup><br>(95% CI) | (3)<br>Number of mail-order prescriptions<br>Mean<sup>a</sup> (95% CI) |
| --- | --- | --- | --- |
| **Overall** | 500217 | 9.8 (9.5, 10.0) | 1.9 (1.8, 2.0) |
| **Socio-demographic variables** |  |  |  |
| Age groups |  |  |  |
| 18-44 | 251902 | 3.3 (3.1, 3.4) | 0.4 (0.4, 0.4) |
| 45-64 | 163275 | 12.3 (11.9, 12.7) | 2.3 (2.2, 2.4) |
| 65+ | 85040 | 23.1 (22.5, 23.8) | 5.4 (5.2, 5.6) |
| Sex |  |  |  |
| Men | 230339 | 8.9 (8.6, 9.1) | 1.7 (1.7, 1.8) |
| Women | 269878 | 10.6 (10.3, 10.9) | 2.1 (2.0, 2.1) |
| Race/ethnicity |  |  |  |
| Non-Hispanic white | 257530 | 12.2 (11.9, 12.6) | 2.4 (2.3, 2.4) |
| Non-Hispanic black | 82928 | 4.8 (4.5, 5.1) | 1.1 (1.0, 1.2) |
| Non-Hispanic Asian | 28798 | 5.8 (5.2, 6.3) | 0.8 (0.7, 0.9) |
| Non-Hispanic others | 8816 | 7.2 (6.2, 8.3) | 2.0 (1.6, 2.3) |
| Hispanic | 122145 | 3.2 (3.0, 3.4) | 0.6 (0.6, 0.7) |
| Educational attainment<sup>b</sup> |  |  |  |
| Less than high school | 115209 | 5.4 (5.1, 5.7) | 1.4 (1.3, 1.5) |
| High school graduate | 161754 | 9.7 (9.4, 10.1) | 2.0 (1.9, 2.1) |
| College or above | 219498 | 11.2 (10.9, 11.6) | 2.0 (1.9, 2.1) |
| Marital status<sup>c</sup> |  |  |  |
| Married | 263451 | 12.2 (11.8, 12.5) | 2.2 (2.1, 2.3) |
| Divorced, widowed, or separated | 102704 | 11.4 (11.0, 11.8) | 2.8 (2.7, 3.0) |
| Never-married | 134047 | 3.5 (3.3, 3.7) | 0.6 (0.5, 0.6) |
| Household poverty level<sup>d</sup> |  |  |  |
| Low-income | 179268 | 6.4 (6.1, 6.6) | 1.7 (1.6, 1.8) |
| Middle-income | 145067 | 8.9 (8.6, 9.2) | 1.8 (1.7, 1.9) |
| High-income | 153863 | 12.7 (12.3, 13.1) | 2.1 (2.0, 2.2) |
| Region of residence<sup>e</sup> |  |  |  |
| Northeast | 81414 | 10.8 (10.2, 11.5) | 1.9 (1.7, 2.0) |
| Midwest | 99250 | 10.7 (10.1, 11.2) | 2.1 (1.9, 2.2) |
| South | 188945 | 9.4 (9.0, 9.8) | 2.0 (1.9, 2.1) |
| West | 130604 | 8.7 (8.3, 9.2) | 1.6 (1.5, 1.7) |
| **Health characteristics and access to care** |  |  |  |
| Self-reported health status<sup>f</sup> |  |  |  |
| Excellent | 70152 | 5.9 (5.6, 6.2) | 0.6 (0.6, 0.7) |
| Very good | 130808 | 10.4 (10.1, 10.8) | 1.5 (1.4, 1.5) |
| Good | 124694 | 12.9 (12.5, 13.3) | 2.7 (2.6, 2.8) |
| Fair | 52871 | 14.0 (13.4, 14.6) | 4.7 (4.4, 4.9) |
| Poor | 12799 | 13.8 (12.8, 14.8) | 6.4 (5.8, 7.0) |
| Frequency pain interfered with work<sup>g</sup> |  |  |  |
| None or a bit of the time | 306581 | 9.7 (9.4, 10.0) | 1.6 (1.5, 1.6) |
| Some, most, or all of the time | 85008 | 15.1 (14.6, 15.6) | 4.6 (4.4, 4.8) |
| Any limitation<sup>h</sup> |  |  |  |
| No | 368106 | 7.8 (7.5, 8.0) | 1.1 (1.0, 1.1) |
| Yes | 126640 | 15.9 (15.5, 16.4) | 4.4 (4.3, 4.6) |
| Number of chronic conditions<sup>i</sup> |  |  |  |
| None | 245346 | 3.1 (3.0, 3.3) | 0.3 (0.3, 0.3) |
| One | 86035 | 11.8 (11.4, 12.2) | 1.5 (1.4, 1.5) |
| Two | 46574 | 19.8 (19.1, 20.4) | 3.5 (3.4, 3.7) |
| Three or more | 53577 | 26.1 (25.3, 26.9) | 8.0 (7.7, 8.3) |
| Has any health insurance |  |  |  |
| Medicare | 97117 | 21.8 (21.3, 22.4) | 5.5 (5.3, 5.7) |
| Medicaid or public insurance only | 62069 | 3.2 (2.9, 3.5) | 0.8 (0.7, 0.9) |
| Private insurance only | 248103 | 8.4 (8.2, 8.7) | 1.2 (1.7, 2.3) |
| Military insurance only | 7591 | 10.5 (9.3, 11.6) | 2.0 (1.2, 1.2) |
| Uninsured | 85337 | 1.6 (1.5, 1.8) | 0.3 (0.2, 0.3) |
| Travel time to usual place of care<sup>j</sup> |  |  |  |
| No usual source of care | 97022 | 2.6 (2.4, 2.8) | 0.4 (0.3, 0.4) |
| < 15 mins | 131966 | 12.7 (12.2, 13.2) | 2.5 (2.4, 2.6) |
| 15-30 mins | 105516 | 14.1 (13.6, 14.6) | 3.0 (2.9, 3.1) |
| 31-60 mins | 24116 | 14.5 (13.6, 15.3) | 3.3 (3.0, 3.5) |
| > 60 mins | 4832 | 14.7 (13.1, 16.3) | 4.0 (3.2, 4.8) |

a: Weighted estimates using sampling weight.
b: Excludes 3756 respondents whose educational attainment was not collected.
c: Excludes 15 respondents whose marital status was not collected.
d: Households were classified into three categories based on their total income and household composition: low-income (less than 200% of the federal poverty threshold), middle-income (200-399%), and high-income ( ≥ 400%).
e: Excludes 4 respondents whose region of residence was not collected.
f: Excludes 108893 respondents whose health status was not collected. MEPS started collecting this variable in 2000. Respondents interviewed before 2000 were removed for this particular variable.
g: Excludes 108628 respondents whose information on pain interference was not collected. MEPS started collecting this variable in 2000. Respondents interviewed before 2000 were removed for this particular variable.
h: Excludes 5471 respondents whose limitation status was not collected. The variable was constructed based on whether a person has any instrumental activities of daily living (IADL) limitations, activities of daily living (ADL) limitations, functional limitations, activity limitations, hearing impairment, or vision impairment.
i: Excludes 68685 respondents whose information on chronic conditions was not collected. In 2016, MEPS started using the International Classification of Diseases 10 (ICD-10) instead of ICD-9. As such, the medical classifications of conditions in 2016 are still under review. Respondents interviewed after 2015 were removed for this particular variable. The following conditions are classified as chronic: hypertension, congestive heart failure, coronary artery disease, cardiac arrhythmias, hyperlipidemia, stroke, arthritis, asthma, cancer, chronic kidney disease, chronic obstructive pulmonary disease, dementia (Alzheimer's and other senile dementias), diabetes, hepatitis, human immunodeficiency virus (HIV), and osteoporosis.

**Appendix Table 2:** Prevalence of mail-order prescription use among U.S. adults for the 100 most commonly mailed medications – 1996-2018.

|  | Percent of all adults who received a mail-order prescription for the medication (95% CI)<sup>a</sup> |
| --- | --- |
|  | N = 499372<sup>a</sup> |
| Acetaminophen-Hydrocodone | 6.3 (6.1, 6.5) |
| Levothyroxine | 6.1 (5.9, 6.2) |
| Lisinopril | 5.8 (5.6, 5.9) |
| Azithromycin | 5.6 (5.5, 5.8) |
| Atorvastatin | 5.2 (5.1, 5.4) |
| Amoxicillin | 5.1 (5.0, 5.2) |
| Simvastatin | 4.7 (4.6, 4.9) |
| Metoprolol | 4.3 (4.2, 4.5) |
| Metformin | 4.2 (4.1, 4.3) |
| Ibuprofen | 4.0 (3.9, 4.1) |
| Albuterol | 3.8 (3.7, 3.9) |
| Amlodipine | 3.6 (3.5, 3.7) |
| Omeprazole | 3.6 (3.5, 3.7) |
| Hydrochlorothiazide | 3.6 (3.5, 3.7) |
| Prednisone | 3.2 (3.1, 3.2) |
| Atenolol | 2.8 (2.7, 2.9) |
| Furosemide | 2.7 (2.7, 2.8) |
| Naproxen | 2.3 (2.2, 2.4) |
| Acetaminophen-Oxycodone | 2.2 (2.1, 2.3) |
| Cephalexin | 2.2 (2.1, 2.2) |
| Cyclobenzaprine | 2.2 (2.1, 2.2) |
| Sertraline | 2.2 (2.1, 2.2) |
| Potassium Chloride | 2.1 (2.1, 2.2) |
| Acetaminophen-Codeine | 2.0 (2.0, 2.1) |
| Fluticasone (nasal) | 2.0 (1.9, 2.1) |
| Gabapentin | 1.8 (1.8, 1.9) |
| Conjugated Estrogens | 1.8 (1.7, 1.9) |
| Alprazolam | 1.8 (1.7, 1.9) |
| Ciprofloxacin | 1.8 (1.7, 1.8) |
| Tramadol | 1.7 (1.7, 1.8) |
| Pravastatin | 1.7 (1.6, 1.7) |
| Losartan | 1.6 (1.6, 1.7) |
| Fluoxetine | 1.6 (1.6, 1.7) |
| Warfarin | 1.6 (1.6, 1.7) |
| Sulfamethoxazole-Trimethoprim | 1.5 (1.5, 1.6) |
| Ranitidine | 1.5 (1.5, 1.6) |
| Bacitracin/Neomycin/Polymyxin B (topical) | 1.5 (1.4, 1.6) |
| Bupropion | 1.5 (1.4, 1.5) |
| Clopidogrel | 1.4 (1.4, 1.5) |
| Aspirin | 1.4 (1.3, 1.5) |
| Citalopram | 1.4 (1.3, 1.5) |
| Zolpidem | 1.4 (1.3, 1.4) |
| Amoxicillin-Clavulanate | 1.4 (1.3, 1.4) |
| Esomeprazole | 1.3 (1.3, 1.4) |
| Pantoprazole | 1.3 (1.3, 1.4) |
| Glipizide | 1.3 (1.3, 1.4) |
| Celecoxib | 1.3 (1.3, 1.4) |
| Fexofenadine | 1.3 (1.2, 1.4) |
| Loratadine | 1.3 (1.2, 1.3) |
| Paroxetine | 1.3 (1.2, 1.3) |
| Hydrochlorothiazide-Triamterene | 1.3 (1.2, 1.3) |
| Montelukast | 1.3 (1.2, 1.3) |
| Escitalopram | 1.3 (1.2, 1.3) |
| Trazodone | 1.2 (1.2, 1.3) |
| Fluticasone-Salmeterol | 1.2 (1.2, 1.3) |
| Doxycycline | 1.2 (1.2, 1.3) |
| Ethinyl Estradiol-Norethindrone | 1.2 (1.2, 1.3) |
| Meloxicam | 1.2 (1.2, 1.3) |
| Diltiazem | 1.2 (1.2, 1.3) |
| Acetaminophen-Propoxyphene | 1.2 (1.1, 1.3) |
| Ethinyl Estradiol-Norgestimate | 1.2 (1.1, 1.3) |
| Rosuvastatin | 1.2 (1.1, 1.3) |
| Lansoprazole | 1.2 (1.1, 1.2) |
| Alendronate | 1.2 (1.1, 1.2) |
| Oxycodone | 1.1 (1.1, 1.2) |
| Lorazepam | 1.1 (1.1, 1.2) |
| Hydrochlorothiazide-Lisinopril | 1.1 (1.0, 1.1) |
| Levofloxacin | 1.1 (1.0, 1.1) |
| Tamsulosin | 1.1 (1.0, 1.1) |
| Cetirizine | 1.0 (1.0, 1.1) |
| Estradiol | 1.0 (1.0, 1.1) |
| Venlafaxine | 1.0 (1.0, 1.1) |
| Clonazepam | 1.0 (1.0, 1.1) |
| Amitriptyline | 1.0 (1.0, 1.0) |
| Homatropine-Hydrocodone | 1.0 (0.9, 1.1) |
| Carvedilol | 1.0 (0.9, 1.1) |
| Digoxin | 1.0 (1.0, 1.0) |
| Methylprednisolone | 1.0 (0.9, 1.0) |
| Lovastatin | 1.0 (0.9, 1.0) |
| Penicillin | 1.0 (0.9, 1.0) |
| Multivitamin, Prenatal | 1.0 (0.9, 1.0) |
| Acetaminophen | 0.9 (0.9, 1.0) |
| Valsartan | 0.9 (0.9, 1.0) |
| Allopurinol | 0.9 (0.9, 1.0) |
| Glyburide | 0.9 (0.9, 1.0) |
| Nitroglycerin | 0.9 (0.9, 1.0) |
| Enalapril | 0.9 (0.8, 0.9) |
| Propranolol | 0.8 (0.8, 0.9) |
| Metronidazole | 0.8 (0.8, 0.9) |
| Insulin Glargine | 0.8 (0.8, 0.9) |
| Ethinyl Estradiol-Levonorgestrel | 0.8 (0.8, 0.9) |
| Verapamil | 0.8 (0.8, 0.8) |
| Nifedipine | 0.8 (0.8, 0.8) |
| Fenofibrate | 0.8 (0.7, 0.8) |
| Diazepam | 0.8 (0.7, 0.8) |
| Clarithromycin | 0.8 (0.7, 0.8) |
| Promethazine | 0.8 (0.7, 0.8) |
| Spironolactone | 0.8 (0.7, 0.8) |
| Benzonatate | 0.7 (0.7, 0.8) |
| Diclofenac | 0.7 (0.7, 0.8) |

a: The numerator was the number of adult MEPS participants who obtained a mail-order prescription for the medication, and the denominator was all adult MEPS participants.
b: Excluded 845 respondents from the analytic sample of 500,217 persons who had missing information on medication name(s).
Abbreviation: CI, confidence interval.

**Appendix Table 3:** Prevalence of mail-order prescription use among adults who have been prescribed each medication, for the 100 medications with the highest prevalence – 1996-2018.

|  | No. of users | Percent of users of the medication who obtained a mail-order prescription (95% CI)<sup>a</sup> |
| --- | --- | --- |
| Ezetimibe | 2420 | 35.9 (33.0, 38.8) |
| Ezetimibe-Simvastatin | 2001 | 34.7 (31.5, 38.0) |
| Raloxifene | 1439 | 34.3 (30.9, 37.6) |
| Hydrochlorothiazide-Irbesartan | 680 | 34.2 (30.5, 37.9) |
| Ibandronate | 615 | 32.6 (29.2, 36.1) |
| Niacin | 1616 | 32.5 (29.6, 35.3) |
| Dutasteride | 552 | 32.3 (28.6, 35.9) |
| Calcitonin | 534 | 31.9 (27.3, 36.4) |
| Alendronate | 5400 | 31.3 (29.4, 33.2) |
| Risedronate | 1403 | 31.1 (28.2, 34.0) |
| Cyclosporine (ophthalmic) | 783 | 30.8 (27.0, 34.6) |
| Calcitriol | 545 | 30.8 (26.6, 35.0) |
| Estradiol (topical) | 951 | 30.7 (27.3, 34.2) |
| Nadolol | 562 | 30.6 (27.4, 33.8) |
| Irbesartan | 1646 | 30.0 (26.9, 33.2) |
| Torsemide | 730 | 29.8 (26.3, 33.4) |
| Pioglitazone | 3405 | 29.8 (26.9, 32.7) |
| Rosuvastatin | 5343 | 29.8 (27.8, 31.8) |
| Desloratadine | 1045 | 29.6 (26.0, 33.3) |
| Omega-3 Polyunsaturated Fatty Acids | 1175 | 29.6 (26.6, 32.7) |
| Fenofibrate | 3506 | 29.5 (27.1, 31.9) |
| Mesalamine | 914 | 29.3 (25.6, 33.0) |
| Esomeprazole | 6500 | 29.3 (27.3, 31.3) |
| Ramipril | 2537 | 29.3 (26.5, 32.0) |
| Azelastine (nasal) | 1093 | 29.2 (25.7, 32.7) |
| Sitagliptin | 1806 | 29.2 (25.8, 32.6) |
| Quinapril | 2555 | 29.1 (26.4, 31.7) |
| Pramipexole | 722 | 28.9 (25.1, 32.7) |
| Hydrochlorothiazide-Valsartan | 2174 | 28.8 (26.1, 31.5) |
| Atorvastatin | 24437 | 28.7 (27.6, 29.8) |
| Brimonidine (ophthalmic) | 1332 | 28.7 (25.4, 31.9) |
| Allopurinol | 4319 | 28.5 (26.3, 30.7) |
| Nebivolol | 775 | 28.4 (24.6, 32.1) |
| Testosterone | 1023 | 28.3 (25.1, 31.5) |
| Apixaban | 565 | 28.2 (24.7, 31.7) |
| Terazosin | 2550 | 28.1 (25.6, 30.5) |
| Tamsulosin | 4738 | 28.0 (26.0, 30.0) |
| Doxazosin | 2324 | 28.0 (25.1, 30.8) |
| Telmisartan | 627 | 27.9 (23.5, 32.3) |
| Levocetirizine | 502 | 27.8 (23.7, 32.0) |
| Latanoprost (ophthalmic) | 2934 | 27.8 (25.5, 30.2) |
| Ropinirole | 929 | 27.7 (23.7, 31.6) |
| Solifenacin | 679 | 27.7 (23.3, 32.1) |
| Insulin Lispro | 1497 | 27.5 (24.5, 30.6) |
| Memantine | 780 | 27.5 (24.0, 31.0) |
| Bimatoprost (ophthalmic) | 883 | 27.4 (24.4, 30.5) |
| Budesonide | 815 | 27.4 (24.3, 30.4) |
| Hydrochlorothiazide-Losartan | 3017 | 27.3 (24.8, 29.9) |
| Rivaroxaban | 627 | 27.3 (24.0, 30.7) |
| Finasteride | 2188 | 27.2 (24.7, 29.8) |
| Tolterodine | 1556 | 27.2 (24.2, 30.1) |
| Azathioprine | 544 | 27.1 (23.1, 31.1) |
| Valsartan | 4628 | 27.0 (24.9, 29.1) |
| Losartan | 8094 | 26.8 (25.2, 28.5) |
| Dorzolamide-Timolol (ophthalmic) | 929 | 26.8 (23.5, 30.1) |
| Clopidogrel | 7003 | 26.7 (25.0, 28.4) |
| Simvastatin | 22737 | 26.7 (25.6, 27.9) |
| Sildenafil | 1089 | 26.5 (22.9, 30.1) |
| Insulin Isophane (Nph) | 1340 | 26.5 (23.0, 29.9) |
| Sulfasalazine | 568 | 26.4 (21.5, 31.3) |
| Olmesartan | 1434 | 26.3 (23.0, 29.7) |
| Insulin Aspart | 1696 | 26.3 (23.3, 29.4) |
| Travoprost (ophthalmic) | 970 | 26.3 (22.7, 29.9) |
| Anastrozole | 587 | 26.2 (22.5, 30.0) |
| Diclofenac-Misoprostol | 656 | 26.2 (23.1, 29.3) |
| Tiotropium | 1922 | 26.0 (23.0, 28.9) |
| Carbidopa-Levodopa | 889 | 25.8 (22.8, 28.7) |
| Hydrochlorothiazide-Olmesartan | 908 | 25.7 (21.5, 29.9) |
| Glimepiride | 3368 | 25.7 (23.0, 28.3) |
| Methotrexate | 1408 | 25.6 (22.7, 28.6) |
| Liraglutide | 500 | 25.6 (21.7, 29.5) |
| Dexlansoprazole | 581 | 25.5 (21.2, 29.9) |
| Donepezil | 1770 | 25.3 (22.4, 28.2) |
| Estradiol | 4334 | 25.3 (23.5, 27.1) |
| Insulin Glargine | 4350 | 25.3 (23.2, 27.4) |
| Polyethylene Glycol 3350 | 801 | 25.2 (21.9, 28.6) |
| Amiodarone | 1129 | 25.2 (22.1, 28.3) |
| Pravastatin | 8199 | 25.1 (23.4, 26.8) |
| Montelukast | 5828 | 25.0 (23.3, 26.6) |
| Celecoxib | 6521 | 24.8 (23.2, 26.4) |
| Tamoxifen | 997 | 24.7 (21.8, 27.6) |
| Rosiglitazone | 1783 | 24.6 (21.4, 27.8) |
| Bumetanide | 683 | 24.6 (21.8, 27.4) |
| Warfarin | 7395 | 24.4 (22.9, 26.0) |
| Carvedilol | 5233 | 24.4 (22.6, 26.3) |
| Bisacodyl | 574 | 24.3 (20.4, 28.2) |
| Isosorbide Mononitrate | 2725 | 24.3 (22.1, 26.5) |
| Atropine-Diphenoxylate | 1045 | 24.3 (21.0, 27.6) |
| Metoprolol | 20708 | 24.3 (23.2, 25.3) |
| Metformin-Sitagliptin | 684 | 24.2 (20.5, 28.0) |
| Amlodipine-Benazepril | 3059 | 24.2 (21.8, 26.7) |
| Levothyroxine | 26878 | 24.2 (23.2, 25.1) |
| Gemfibrozil | 2550 | 24.0 (21.7, 26.3) |
| Diclofenac (topical) | 967 | 24.0 (20.9, 27.0) |
| Timolol (ophthalmic) | 2086 | 23.9 (21.3, 26.6) |
| Hydroxychloroquine | 1475 | 23.9 (20.9, 26.8) |
| Eszopiclone | 633 | 23.8 (20.1, 27.5) |
| Diltiazem | 5886 | 23.8 (22.2, 25.5) |
| Insulin Detemir | 1035 | 23.8 (20.0, 27.6) |
| Indapamide | 715 | 23.7 (20.3, 27.2) |

a: The numerator was the number of adult MEPS participants who obtained a mail-order prescription for the medication, and the denominator was the number of adult MEPS participants who have been prescribed the medication.
Abbreviation: CI, confidence interval.

## References

1 DeSilver D. The state of the U.S. Postal Service in 8 charts. https://www.pewresearch.org/facttank/2020/05/14/the-state-of-the-u-s-postal-service-in-8-charts/ (accessed Aug 21, 2020).

2 Statement by NALC President Fredric Rolando: The Postal Service is vital in this crisis. The National Association of Letter Carriers. 2020; published online March 27. https://www.nalc.org/news/nalc-updates/statement-by-nalc-president-fredric-rolando-the-postal-service-is-vital-in-this-crisis (accessed Aug 23, 2020).

3 Broadwater L, Healy J, Shear M, Fuchs H. Postal Crisis Ripples Across Nation as Election Looms. The New York Times. 2020; published online Aug 15. https://www.nytimes.com/2020/08/15/us/post-office-vote-by-mail.html (accessed Aug 22, 2020).

4 Baker M. The Facts About Mail-In Voting and Voter Fraud. The New York Times. 2020; published online Aug 26. https://www.nytimes.com/article/mail-in-vote-fraud-ballot.html (accessed Aug 31, 2020).

5 Carroll NV, Brusilovsky I, York B, Oscar R. Comparison of Costs of Community and Mail Service Pharmacy. Journal of the American Pharmacists Association 2005; 45: 336–43.

6 Schmittdiel JA, Karter AJ, Dyer W, et al. The Comparative Effectiveness of Mail Order Pharmacy Use vs. Local Pharmacy Use on LDL-C Control in New Statin Users. Journal of General Internal Medicine 2011; 26: 1396–402.

7 Devine S, Vlahiotis A, Sundar H. A comparison of diabetes medication adherence and healthcare costs in patients using mail order pharmacy and retail pharmacy. Journal of Medical Economics 2010; 13: 203–11.

8 Jackson C, Newall M. Between Democratic and Republican conventions, Americans more likely to trust Biden on COVID-19 than Trump. 2020; published online Aug 24. https://www.ipsos.com/en-us/news-polls/axios-ipsos-coronavirus-index (accessed Aug 27, 2020).

9 Axios/Ipsos Poll – Wave 22. Axios, 2020 https://www.ipsos.com/sites/default/files/ct/news/documents/2020-08/topline-axios-w22.pdf (accessed Aug 29, 2020).

10 Hopkins J. Mail-Order Drug Delivery Rises During Coronavirus Lockdowns. 2020; published online May 12. https://www.wsj.com/articles/mail-order-drug-delivery-rises-during-coronavirus-lockdowns-11589281203 (accessed Aug 27, 2020).

11 Sen-Crowe B, McKenney M, Elkbuli A. Medication shortages during the COVID-19 pandemic: Saving more than COVID lives. The American Journal of Emergency Medicine DOI:10.1016/j.ajem.2020.07.044.

12 Ma J, Wang L. Characteristics of Mail-Order Pharmacy Users: Results From the Medical Expenditures Panel Survey. Journal of Pharmacy Practice 2018; 33: 293–8.

13 Phelan J, Ergun D, Langer G, Holyk G. Medication Adherence in America: A National Report Card. National Community Pharmacists Association, 2013 http://www.ncpa.co/adherence/AdherenceReportCard_Full.pdf (accessed Aug 22, 2020).

14 Cubanski J, Biniek J, Rae M, Damico A, Frederiksen B, Salganicoff A. Mail Delays Could Affect Mail-Order Prescriptions for Millions of Medicare Part D and Large Employer Plan Enrollees. 2020 https://www.kff.org/coronavirus-covid-19/issue-brief/mail-delays-could-affect-mail-order-prescriptions-for-millions-of-medicare-part-d-and-large-employer-plan-enrollees/ (accessed Aug 29, 2020).

15 Rashrash ME, Tomaszewski DM, Schommer JC, Brown LM. Consumers’ characteristics associated with the use of mail pharmacy services in the United States: Findings from the 2015 National Consumer Survey on the Medication Experience. Journal of the American Pharmacists Association 2017; 57: 206–10.

16 Medicine Spending and Affordability in the United States. IQVIA Institute for Human Data Science, 2020 https://www.iqvia.com/insights/the-iqvia-institute/reports/medicine-spending-and-affordability-in-the-us?fbclid=IwAR1yC55Ib1CHHXimCe7rJdxwDD1q2yKt9fdwE1IeXJ8gqDp5rqbnTuMHaMo (accessed Aug 22, 2020).

17 Blewett LA, Drew JAR, Griffin R, Williams KC. IPUMS Health Surveys: Medical Expenditure Panel Survey, Version 1.1 [dataset]. Minneapolis: University of Minnesota 2019. https://meps.ipums.org/ (accessed Aug 23, 2020).

18 Agency for healthcare Research and Quality. MEPS-HC Response Rates by Panel. 2019; published online Aug 30. https://meps.ahrq.gov/survey_comp/hc_response_rate.jsp (accessed Aug 25, 2020).

19 MEPS Household Component. Annual Contractor Methodology Report 2017. Agency for Healthcare Research and Quality, 2018 https://meps.ahrq.gov/data_files/publications/annual_contractor_report/hc_ann_cntrct_methrpt.shtml (accessed Aug 30, 2020).

20 Hill SC, Zuvekas SH, Zodet MW. Implications of the accuracy of MEPS prescription drug data for health services research. Inquiry 2011; 48: 242–59.

21 Goodman RA, Posner SF, Huang ES, Parekh AK, Koh HK. Defining and measuring chronic conditions: imperatives for research, policy, program, and practice. Prev Chronic Dis 2013; 10: E66.

22 Profiles of General Demographic Characteristics. 2000 Census of Population and Housing. U.S. Census Bureau, 2001 https://www.census.gov/prod/cen2000/dp1/2khus.pdf.(accessed Aug 30, 2020).

23 Rupp MT. Attitudes of Medicare-Eligible Americans Toward Mail Service Pharmacy. JMCP 2013; 19: 564–72.

24 Pub. 100-18 Medicare Prescription Drug Benefit Manual. Department of Health & Human Services. Center for medicare & Medicaid Services, 2008 https://www.cms.gov/Regulations-and-Guidance/Guidance/Transmittals/downloads/R1PDB.pdf (accessed Aug 27, 2020).

25 Wu J, Davis–Ajami ML, Noxon V. Patterns of use and expenses associated with mail-service pharmacy in adults with diabetes. Journal of the American Pharmacists Association 2015; 55: 41–51.

26 Williamson EJ, Walker AJ, Bhaskaran K, et al. Factors associated with COVID-19-related death using OpenSAFELY. Nature 2020; 584: 430–6.

27 Mantwill S, Monestel-Umaña S, Schulz PJ. The Relationship between Health Literacy and Health Disparities: A Systematic Review. PLOS ONE 2015; 10: e0145455.

28 Centers for Disease Control and Prevention (CDC). National Health and Nutrition Examination Survey. https://www.cdc.gov/nchs/nhanes/index.htm.

29 Behavioral Risk Factor Surveillance System. 2018 Summary Data Quality Report. Centers for Disease Control and Prevention (CDC), 2019 https://www.cdc.gov/brfss/annual_data/2018/pdf/2018-sdqr-508.pdf (accessed Sept 1, 2020).

30 Abelson R. U.S. Mail Delays Slow Delivery of Medicines. New York Times. 2020; published online Aug 20. https://www.nytimes.com/2020/08/20/health/Covid-us-mail-prescription-drugs.html (accessed Aug 22, 2020).

